# A Three-Tier Operational Benchmark for Evaluating Large Language Models on Hospital Medication Safety

**DOI:** 10.64898/2026.06.05.26354271

**Authors:** Joshua Proulx, Bryce Daines, Michael Barton, Molly E. Leonard, Joseph A. Garcia, Bronson Young, Quinn Snell, Timothy W. West, Sam R. Watson, Maryam AlQaseer, Mathieu Louiset, Muhammad Bilal Maqsood, Mary J. Voutt-Goos, Caryn Douma, Nishaminy Kasbekar, Jaclyn Jeffries, Wadie Abu-Rahmeh, Karen Frush, Darshan K. Grewal, Mouna Bahsoun, Michael Leonard, Allan Frankel, David C. Classen, Stanley L. Pestotnik

## Abstract

**Objective:** To introduce PsiBench, a clinically validated medication-safety benchmark for evaluating large language models (LLMs) against the standards used to certify hospital computerized provider order entry (CPOE) and electronic health record (EHR) systems, and a non-overlapping three-tier evaluation framework separating highest-stakes discrimination, the operational CDS regime, and category-correct alerting.

**Materials and Methods:** PsiBench comprises 492 medication-safety scenarios across 11 safety categories, created by clinical pharmacology experts whose work underpins an annualized testing procedure used by more than 2,000 U.S. hospitals. The three-tier framework partitions the scenarios non-overlappingly: Discrimination (98 scenarios, 50 fatal vs 48 deception, near-balanced 51%/49%); Operational (394 scenarios, 261 serious unsafe plus 133 safe including 41 Excessive Alerts reclassified as operational negatives); and Attribution (311 alert-required scenarios). We evaluated 40 frontier LLMs from 10 providers over 3 runs per scenario at temperature 0.2 (or the provider default where temperature is not configurable), yielding 59,040 evaluations conducted April 21–23, 2026.

**Results:** Headline binary performance on the full benchmark spans a wide range across the 40 models: F1 78.5%–92.3%, accuracy 65.4%–89.8%, sensitivity 81.4%–100.0%, specificity 6.1%–81.8%. Leading models by F1 (o4-mini 92.3%; o3 92.2%) pair high sensitivity with meaningful specificity; three models saturate sensitivity at 100% but fall below 25% specificity, indistinguishable from a naive always-alert classifier. The wide spread on a single headline metric motivates tier-specific analyses, developed in a separate clinical paper.

**Discussion and Conclusion:** PsiBench and the three-tier framework operationalize a rigorous evaluation rubric for LLM medication safety, grounded in two decades of national hospital audit experience. The framework generalizes to any binary medication-safety classifier (rule-based, conventional ML, or LLM-driven), supporting tier-aware model selection and post-deployment surveillance.

## Introduction

Medication errors remain among the most common and preventable causes of patient harm in hospital settings.^1,2^ In the United States, adverse drug events account for over 700,000 emergency department visits annually,^3^ and are responsible for nearly 100,000 emergency hospitalizations among older adults each year;^4^ published estimates of the annual cost of treating preventable adverse drug events in U.S. hospitals span roughly $0.9–5.1 billion, depending on the medication classes and error types examined.^5–9^ Landmark studies have demonstrated that adverse drug events significantly increase length of stay, healthcare costs, and attributable mortality among hospitalized patients.^10,11^ Computerized provider order entry (CPOE) and electronic health record (EHR) systems equipped with clinical decision support (CDS) represent the primary technological response to this challenge, with systematic reviews confirming their capacity to reduce medication errors when properly implemented.^12,13^ Standardized frameworks for evaluating these systems have been in development since the early 2000s,^14–17^ a period over which CPOE/EHR adoption was also broadly incentivized through national health-information-technology policy initiatives such as Meaningful Use and the 21st Century Cures Act.

Yet two decades after CPOE/EHR-based CDS became standard, hospital medication safety remains uneven across institutions and alert categories, both in catch rates for genuinely fatal orders and in alert volumes that exceed what clinicians can act on.^14–19,40^ Into this baseline, LLM-driven medication-management systems are entering production in U.S. clinical settings; Doctronic’s State of Utah partnership, announced in January 2026, uses an LLM to autonomously process renewals for roughly 190 chronic medications with physician review built into the workflow.^38,39^ The clinical-deployment consequences of that unevenness are taken up in a separate clinical paper; here we focus on the methodological problem it creates: whether and how LLMs can improve on the existing baseline depends entirely on how their performance is measured.

The emergence of large language models (LLMs) as general-purpose reasoning engines has prompted rapid exploration of their potential in clinical applications, including medical question answering, diagnostic reasoning, and drug interaction assessment.^20–23^ A recent scoping review identified three primary areas where generative AI and LLMs are being applied to mitigate medication-related harm: drug-drug interaction identification, clinical decision support, and pharmacovigilance.^24^ Individual studies have demonstrated that LLMs can detect medication direction errors in pharmacy workflows,^25^ identify drug-drug interactions with varying sensitivity and specificity relative to conventional clinical tools,^26,27^ and perform medication safety reviews in primary care settings with high sensitivity but limited full correctness.^28^ Emerging work has benchmarked LLMs against clinical pharmacists in prescription review tasks.^29^ However, these evaluations share critical limitations as a class: they typically assess models on isolated sub-tasks rather than comprehensive ordering workflows, employ small or synthetic datasets without established clinical provenance, and test only one or two models, precluding conclusions about the broader landscape of LLM capabilities. Equally important, most rely on a single aggregate accuracy or F1-type metric computed on a class-imbalanced scenario set, an evaluation choice known to obscure systematic differences in sensitivity-specificity tradeoffs and to inflate apparent performance on the dominant class.^35–37^

We introduce PsiBench, a benchmark designed to bridge this gap. PsiBench comprises 492 clinically validated medication safety scenarios, created by clinical pharmacology experts with decades of experience in hospital medication safety evaluation. This expertise underpins an annualized testing procedure used by more than 2,000 U.S. hospitals to evaluate their CPOE/EHR systems against federally recognized safety criteria.^40^ The benchmark spans 11 safety categories and 3 subcategories, covering drug-allergy contraindications, dose-range violations, drug-drug interactions, renal dosing adjustments, therapeutic duplication, and other safety domains. Critically, unsafe orders in the benchmark are defined with a high bar: these are medication orders known to cause fatal or severe patient harm, not merely potentially unsafe or theoretically problematic prescriptions.

Beyond the benchmark itself, we propose a non-overlapping three-tier evaluation framework that separates three fundamentally different questions about model capability. The **Discrimination tier** tests highest-stakes discrimination on a tight 98-scenario near-balanced subset comprising 50 fatal scenarios and 48 deception scenarios. Deception scenarios are safe medication orders intentionally constructed by clinical pharmacology experts to resemble unsafe orders at first glance, with surface-level alarm features that resolve under closer reading to a clinically appropriate order. They serve as an adversarial specificity test: a model that alerts on a deception scenario has been fooled by surface features rather than reasoning through the clinical context. The **Operational tier** is the operational CDS regime: 394 scenarios comprising 261 serious unsafe orders and 133 safe orders (92 regular safe orders plus 41 Excessive Alerts scenarios reclassified as operational negatives). The **Attribution tier** assesses category-correct alerting across the 311 alert-required scenarios, measuring whether models alert for the correct clinical reason. The Discrimination and Operational tiers are non-overlapping and together account for all 492 benchmark scenarios; the Attribution tier evaluates an orthogonal dimension.

In this paper we describe the benchmark composition, the three-tier evaluation framework, the evaluation protocol used to generate the data, and the summary headline performance of 40 frontier LLMs from 10 providers (59,040 total evaluations conducted April 21–23, 2026). We argue, with empirical demonstration on the headline numbers, that any subsequent operational claim about LLM medication-safety capability should be reported at the tier level rather than as a single aggregate metric, and that the three-tier decomposition is the minimum structure required to make such reports interpretable for deployment decisions. The detailed per-tier empirical findings, including the tier-specific ranking shifts that motivate tier-aware model selection in clinical deployment, are reported in a separate clinical paper.

## Results

### Empirical demonstration that single-metric evaluation hides clinically relevant variation

We instantiate the framework on a 492-scenario benchmark and 40 frontier LLMs evaluated three times each, yielding 59,040 evaluations between April 21 and 23, 2026. The methodological claim of this paper, that single-metric evaluation collapses three clinically distinct questions into one number and obscures large per-tier capability differences, can be falsified directly on the resulting per-model numbers. Two empirical patterns establish that the collapse occurs.

First, the spread on each single binary metric, computed over the full benchmark, is wide enough that the metric cannot in principle convey the rank order it would have under any of the three tier-specific evaluations. Across the 40 models we observe sensitivity ranging from 81.4% to 100.0%, specificity from 6.1% to 81.8%, and F1 from 78.5% to 92.3% (Figure M2; per-model values in Table 1, with operating characteristics in Table 2). The sensitivity ceiling is a trivial consequence of permitting always-alert behavior, and at the saturated end multiple models reach 100% sensitivity with specificity below 25%, indistinguishable from a naive always-alert classifier on a single-metric report. The wide specificity floor is the operationally meaningful tail.

**Figure M1.**
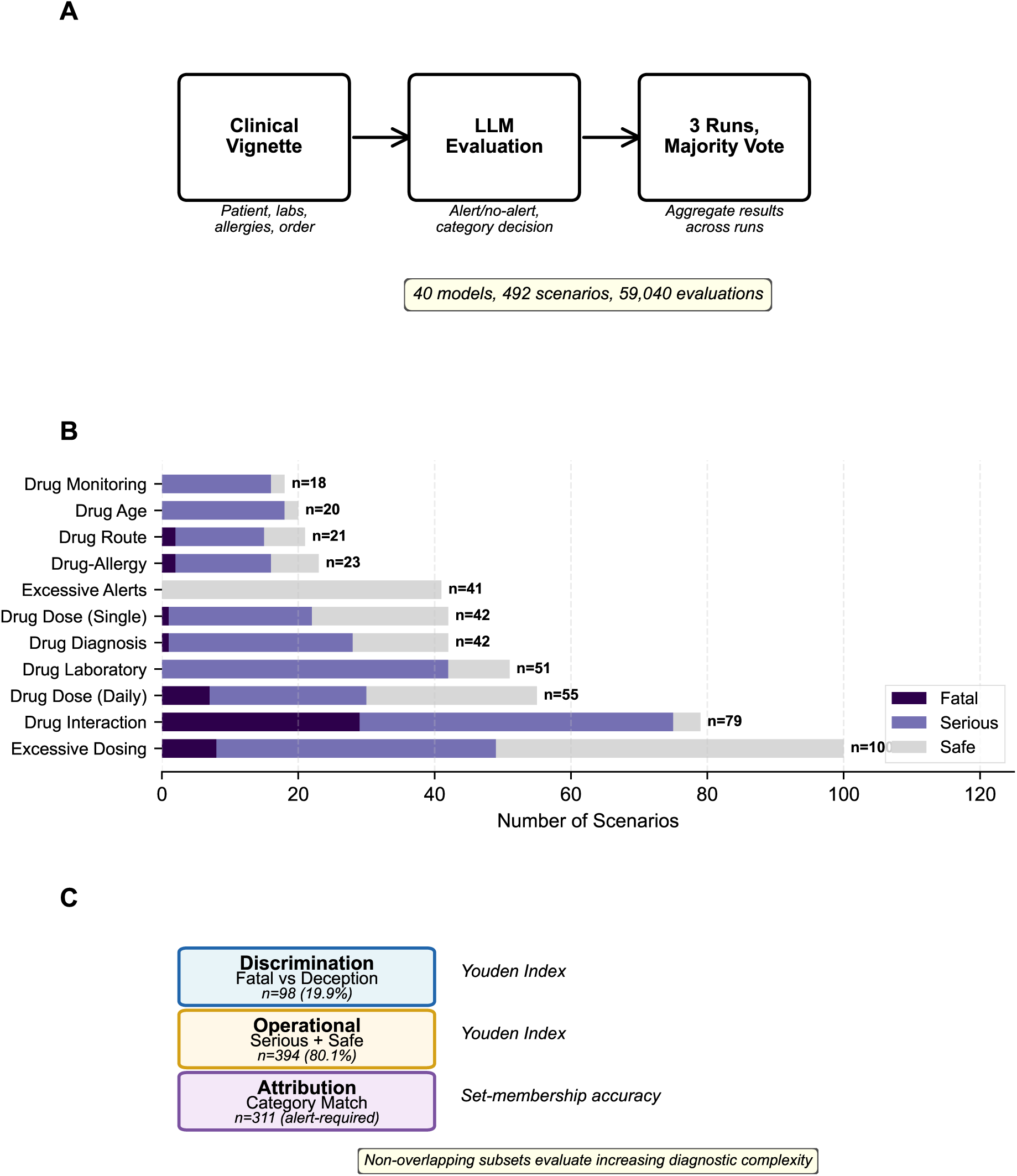
PsiBench and the non-overlapping three-tier evaluation framework. (A) Evaluation pipeline. Each scenario presents a structured patient vignette (demographics, diagnoses, laboratory values, allergies, and a medication order) to the model, which returns an alert decision and a safety-category classification. Each scenario is evaluated three times per model at temperature 0.2 (or the provider default where temperature is not configurable); majority vote across runs determines the final prediction. The benchmark comprises 40 models, 492 scenarios, and 59,040 evaluations performed April 21–23, 2026. (B) Scenario distribution across the 11 safety categories and 3 subcategories (Drug-Drug Interaction, Therapeutic Duplication, Renal), stratified by clinical severity: 50 fatal, 261 serious, and 181 safe (133 regular safe and 48 deception). (C) Three-tier framework. Discrimination tier: 98 scenarios (50 fatal and 48 deception), near-balanced 51%/49%, evaluated by Youden’s J. Operational tier: 394 scenarios (261 serious unsafe and 133 safe, the latter including 41 Excessive Alerts reclassified as operational negatives), 66% alert / 34% safe, evaluated by Youden’s J. Attribution tier: 311 alert-required scenarios (50 fatal and 261 serious), evaluated by category-match rate using set-membership matching. The Discrimination and Operational tiers are non-overlapping and together cover all 492 scenarios; the Attribution tier is orthogonal.

**Figure M2.**
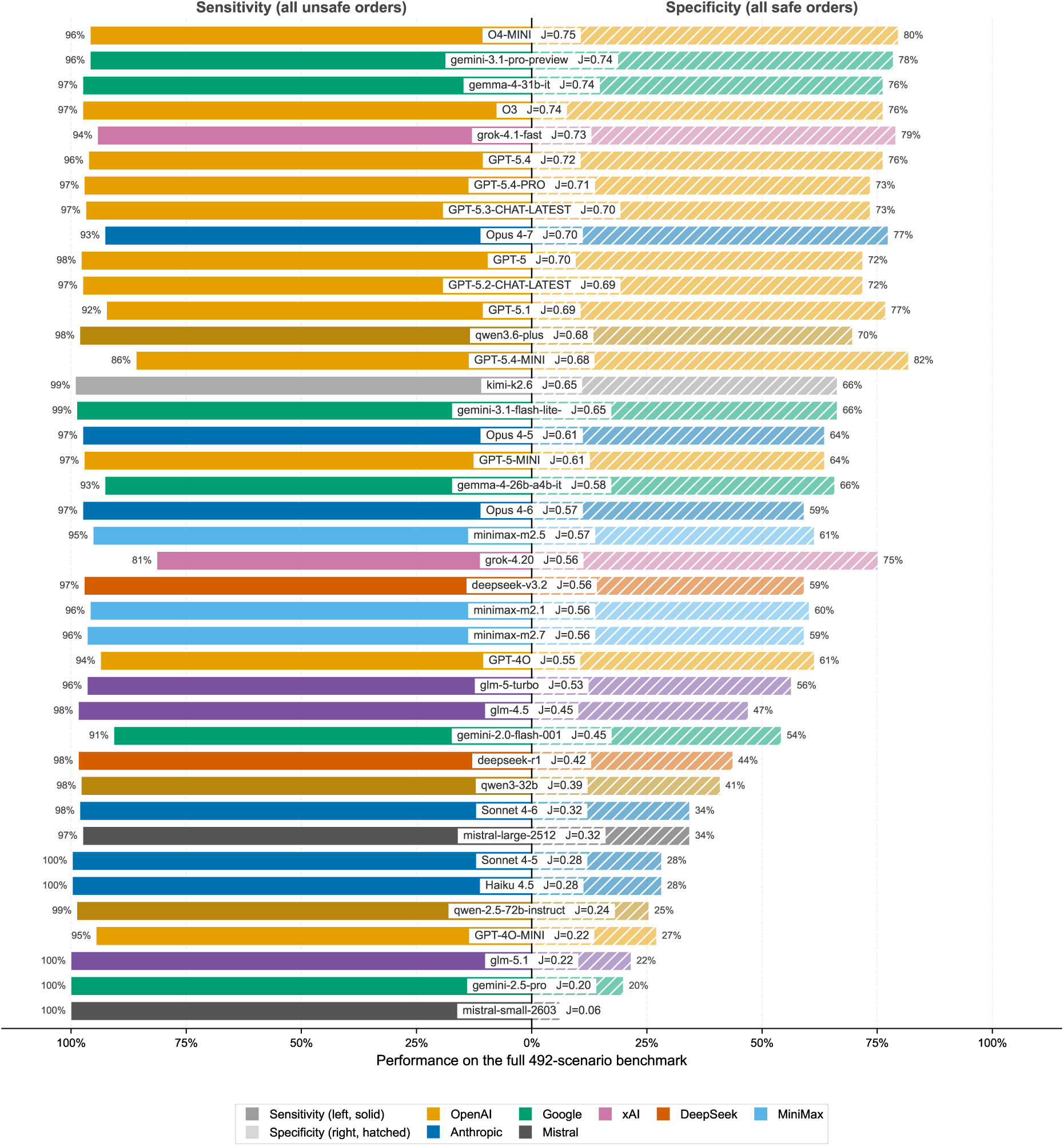
Per-model sensitivity versus specificity on the full 492-scenario benchmark. Butterfly chart showing sensitivity on all unsafe orders (left, solid bars) and specificity on all safe orders (right, hatched bars) for each of the 40 evaluated models. Bars are colored by provider; the same color appears on both sides for a given model. The central column shows the model name plus its Youden’s J value (sensitivity + specificity − 1), with models sorted top-to-bottom by Youden’s J descending. Sensitivity spans 81.4% to 100.0%, specificity 6.1% to 81.8%, and Youden’s J 0.06 to 0.75, exposing the sensitivity-specificity tradeoff that single-metric reporting hides and motivating the three-tier framework introduced in the Methods section. Per-model confusion-matrix counts (TP, FP, TN, FN), sensitivity, specificity, and accuracy are reported in Table 1; precision, NPV, F1, Youden’s J, false-positive rate, alert rate, and median response time are reported in Table 2.

**Table 1.**
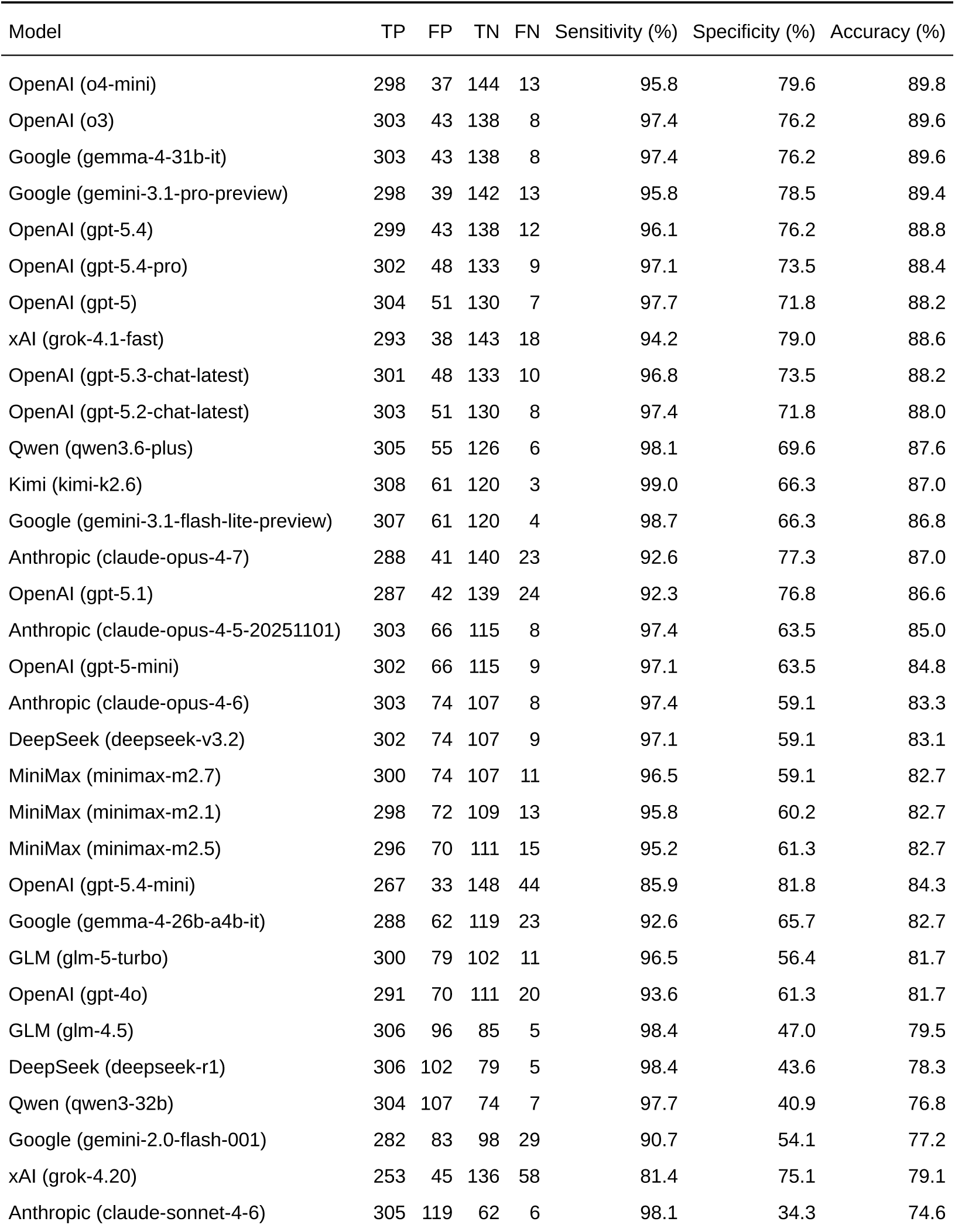
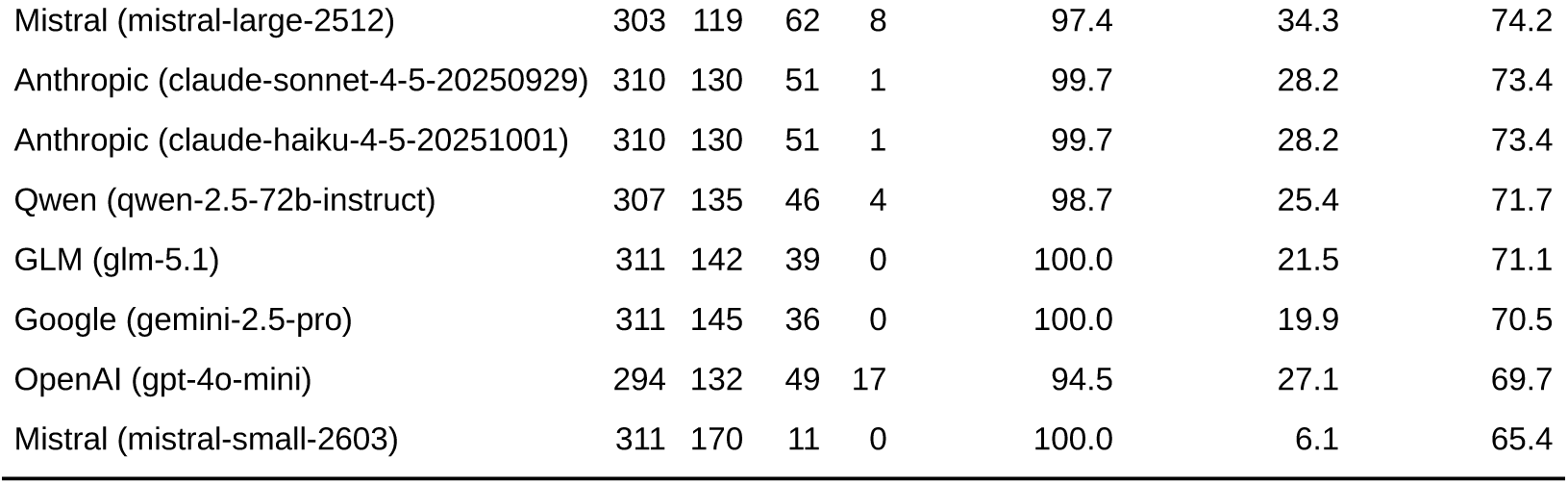
Per-model confusion-matrix counts and primary rate metrics for all 40 models on the full 492-scenario benchmark. Per-model confusion-matrix counts and primary rate metrics on the full 492-scenario benchmark, sorted by F1 score (descending). Computed via majority vote across 3 independent runs per scenario. TP = true positives (correctly alerted unsafe orders); FP = false positives (alerts on safe orders); TN = true negatives (correctly silent on safe orders); FN = false negatives (missed un-safe orders). Sensitivity = TP / (TP + FN). Specificity = TN / (TN + FP). Accuracy = (TP + TN) / total scenarios. Precision, NPV, F1, Youden’s J, false-positive rate, alert rate, and median response time per model are reported in Table 2. Two pairs of models (o3 and gemma-4-31b-it; claude-sonnet-4-5-20250929 and claude-haiku-4-5-20251001) have identical aggregate confusion-matrix counts but differ at the per-scenario level.

| Model | TP | FP | TN | FN | Sensitivity (%) | Specificity (%) | Accuracy (%) |
| --- | --- | --- | --- | --- | --- | --- | --- |
| OpenAI (o4-mini) | 298 | 37 | 144 | 13 | 95.8 | 79.6 | 89.8 |
| OpenAI (o3) | 303 | 43 | 138 | 8 | 97.4 | 76.2 | 89.6 |
| Google (gemma-4-31b-it) | 303 | 43 | 138 | 8 | 97.4 | 76.2 | 89.6 |
| Google (gemini-3.1-pro-preview) | 298 | 39 | 142 | 13 | 95.8 | 78.5 | 89.4 |
| OpenAI (gpt-5.4) | 299 | 43 | 138 | 12 | 96.1 | 76.2 | 88.8 |
| OpenAI (gpt-5.4-pro) | 302 | 48 | 133 | 9 | 97.1 | 73.5 | 88.4 |
| OpenAI (gpt-5) | 304 | 51 | 130 | 7 | 97.7 | 71.8 | 88.2 |
| xAI (grok-4.1-fast) | 293 | 38 | 143 | 18 | 94.2 | 79.0 | 88.6 |
| OpenAI (gpt-5.3-chat-latest) | 301 | 48 | 133 | 10 | 96.8 | 73.5 | 88.2 |
| OpenAI (gpt-5.2-chat-latest) | 303 | 51 | 130 | 8 | 97.4 | 71.8 | 88.0 |
| Qwen (qwen3.6-plus) | 305 | 55 | 126 | 6 | 98.1 | 69.6 | 87.6 |
| Kimi (kimi-k2.6) | 308 | 61 | 120 | 3 | 99.0 | 66.3 | 87.0 |
| Google (gemini-3.1-flash-lite-preview) | 307 | 61 | 120 | 4 | 98.7 | 66.3 | 86.8 |
| Anthropic (claude-opus-4-7) | 288 | 41 | 140 | 23 | 92.6 | 77.3 | 87.0 |
| OpenAI (gpt-5.1) | 287 | 42 | 139 | 24 | 92.3 | 76.8 | 86.6 |
| Anthropic (claude-opus-4-5-20251101) | 303 | 66 | 115 | 8 | 97.4 | 63.5 | 85.0 |
| OpenAI (gpt-5-mini) | 302 | 66 | 115 | 9 | 97.1 | 63.5 | 84.8 |
| Anthropic (claude-opus-4-6) | 303 | 74 | 107 | 8 | 97.4 | 59.1 | 83.3 |
| DeepSeek (deepseek-v3.2) | 302 | 74 | 107 | 9 | 97.1 | 59.1 | 83.1 |
| MiniMax (minimax-m2.7) | 300 | 74 | 107 | 11 | 96.5 | 59.1 | 82.7 |
| MiniMax (minimax-m2.1) | 298 | 72 | 109 | 13 | 95.8 | 60.2 | 82.7 |
| MiniMax (minimax-m2.5) | 296 | 70 | 111 | 15 | 95.2 | 61.3 | 82.7 |
| OpenAI (gpt-5.4-mini) | 267 | 33 | 148 | 44 | 85.9 | 81.8 | 84.3 |
| Google (gemma-4-26b-a4b-it) | 288 | 62 | 119 | 23 | 92.6 | 65.7 | 82.7 |
| GLM (glm-5-turbo) | 300 | 79 | 102 | 11 | 96.5 | 56.4 | 81.7 |
| OpenAI (gpt-4o) | 291 | 70 | 111 | 20 | 93.6 | 61.3 | 81.7 |
| GLM (glm-4.5) | 306 | 96 | 85 | 5 | 98.4 | 47.0 | 79.5 |
| DeepSeek (deepseek-r1) | 306 | 102 | 79 | 5 | 98.4 | 43.6 | 78.3 |
| Qwen (qwen3-32b) | 304 | 107 | 74 | 7 | 97.7 | 40.9 | 76.8 |
| Google (gemini-2.0-flash-001) | 282 | 83 | 98 | 29 | 90.7 | 54.1 | 77.2 |
| xAI (grok-4.20) | 253 | 45 | 136 | 58 | 81.4 | 75.1 | 79.1 |
| Anthropic (claude-sonnet-4-6) | 305 | 119 | 62 | 6 | 98.1 | 34.3 | 74.6 |

Table 1. Per-model confusion-matrix counts and primary rate metrics for all 40 models on the full 492-scenario benchmark.
| Model | TP | FP | TN | FN | Sensitivity (%) | Specificity (%) | Accuracy (%) |
| --- | --- | --- | --- | --- | --- | --- | --- |
| Mistral (mistral-large-2512) | 303 | 119 | 62 | 8 | 97.4 | 34.3 | 74.2 |
| Anthropic (claude-sonnet-4-5-20250929) | 310 | 130 | 51 | 1 | 99.7 | 28.2 | 73.4 |
| Anthropic (claude-haiku-4-5-20251001) | 310 | 130 | 51 | 1 | 99.7 | 28.2 | 73.4 |
| Qwen (qwen-2.5-72b-instruct) | 307 | 135 | 46 | 4 | 98.7 | 25.4 | 71.7 |
| GLM (glm-5.1) | 311 | 142 | 39 | 0 | 100.0 | 21.5 | 71.1 |
| Google (gemini-2.5-pro) | 311 | 145 | 36 | 0 | 100.0 | 19.9 | 70.5 |
| OpenAI (gpt-4o-mini) | 294 | 132 | 49 | 17 | 94.5 | 27.1 | 69.7 |
| Mistral (mistral-small-2603) | 311 | 170 | 11 | 0 | 100.0 | 6.1 | 65.4 |

**Table 2.** Per-model derived metrics and operational profile for all 40 models on the full 492-scenario benchmark. Per-model derived metrics and operational profile on the full 492-scenario benchmark. Rows are sorted by F1 score descending to match the row order in Table 1. Precision = TP / (TP + FP); NPV = TN / (TN + FN); F1 = 2 *·* Precision *·* Sensitivity / (Precision + Sensitivity); Youden’s J = Sensitivity + Specificity *−* 1; FPR = 1 *−* Specificity; Alert Rate = (TP + FP) / total scenarios. Median RT is the median wall-clock API latency per evaluation, including network and provider-side queuing. Confusion-matrix counts and primary rate metrics per model are reported in Table 1.

| Model | Precision (%) | NPV (%) | F1 (%) | Youden's J | FPR (%) | Alert Rate (%) | Median RT (s) |
| --- | --- | --- | --- | --- | --- | --- | --- |
| OpenAI (o4-mini) | 89.0 | 91.7 | 92.3 | 0.754 | 20.4 | 68.1 | 6.3 |
| OpenAI (o3) | 87.6 | 94.5 | 92.2 | 0.737 | 23.8 | 70.3 | 9.9 |
| Google (gemma-4-31b-it) | 87.6 | 94.5 | 92.2 | 0.737 | 23.8 | 70.3 | 6.5 |
| Google (gemini-3.1-pro-preview) | 88.4 | 91.6 | 92.0 | 0.743 | 21.5 | 68.5 | 12.4 |
| OpenAI (gpt-5.4) | 87.4 | 92.0 | 91.6 | 0.724 | 23.8 | 69.5 | 2.3 |
| OpenAI (gpt-5.4-pro) | 86.3 | 93.7 | 91.4 | 0.706 | 26.5 | 71.1 | 133.9 |
| OpenAI (gpt-5) | 85.6 | 94.9 | 91.3 | 0.696 | 28.2 | 72.2 | 21.0 |
| xAI (grok-4.1-fast) | 88.5 | 88.8 | 91.3 | 0.732 | 21.0 | 67.3 | 12.3 |
| OpenAI (gpt-5.3-chat-latest) | 86.2 | 93.0 | 91.2 | 0.703 | 26.5 | 70.9 | 4.2 |
| OpenAI (gpt-5.2-chat-latest) | 85.6 | 94.2 | 91.1 | 0.693 | 28.2 | 72.0 | 5.5 |
| Qwen (qwen3.6-plus) | 84.7 | 95.5 | 90.9 | 0.677 | 30.4 | 73.2 | 48.8 |
| Kimi (kimi-k2.6) | 83.5 | 97.6 | 90.6 | 0.653 | 33.7 | 75.0 | 122.8 |
| Google (gemini-3.1-flash-lite-preview) | 83.4 | 96.8 | 90.4 | 0.650 | 33.7 | 74.8 | 15.7 |
| Anthropic (claude-opus-4-7) | 87.5 | 85.9 | 90.0 | 0.700 | 22.7 | 66.9 | 19.8 |
| OpenAI (gpt-5.1) | 87.2 | 85.3 | 89.7 | 0.691 | 23.2 | 66.9 | 2.9 |
| Anthropic (claude-opus-4-5-20251101) | 82.1 | 93.5 | 89.1 | 0.610 | 36.5 | 75.0 | 38.9 |
| OpenAI (gpt-5-mini) | 82.1 | 92.7 | 89.0 | 0.606 | 36.5 | 74.8 | 16.2 |
| Anthropic (claude-opus-4-6) | 80.4 | 93.0 | 88.1 | 0.565 | 40.9 | 76.6 | 36.7 |
| DeepSeek (deepseek-v3.2) | 80.3 | 92.2 | 87.9 | 0.562 | 40.9 | 76.4 | 5.1 |
| MiniMax (minimax-m2.7) | 80.2 | 90.7 | 87.6 | 0.556 | 40.9 | 76.0 | 18.8 |
| MiniMax (minimax-m2.1) | 80.5 | 89.3 | 87.5 | 0.560 | 39.8 | 75.2 | 15.0 |
| MiniMax (minimax-m2.5) | 80.9 | 88.1 | 87.4 | 0.565 | 38.7 | 74.4 | 13.4 |
| OpenAI (gpt-5.4-mini) | 89.0 | 77.1 | 87.4 | 0.676 | 18.2 | 61.0 | 1.7 |
| Google (gemma-4-26b-a4b-it) | 82.3 | 83.8 | 87.1 | 0.584 | 34.3 | 71.1 | 3.1 |
| GLM (glm-5-turbo) | 79.2 | 90.3 | 87.0 | 0.528 | 43.6 | 77.0 | 21.4 |
| OpenAI (gpt-4o) | 80.6 | 84.7 | 86.6 | 0.549 | 38.7 | 73.4 | 1.5 |
| GLM (glm-4.5) | 76.1 | 94.4 | 85.8 | 0.454 | 53.0 | 81.7 | 29.7 |
| DeepSeek (deepseek-r1) | 75.0 | 94.0 | 85.1 | 0.420 | 56.4 | 82.9 | 76.2 |
| Qwen (qwen3-32b) | 74.0 | 91.4 | 84.2 | 0.386 | 59.1 | 83.5 | 21.4 |
| Google (gemini-2.0-flash-001) | 77.3 | 77.2 | 83.4 | 0.448 | 45.9 | 74.2 | 1.6 |
| xAI (grok-4.20) | 84.9 | 70.1 | 83.1 | 0.565 | 24.9 | 60.6 | 1.3 |
| Anthropic (claude-sonnet-4-6) | 71.9 | 91.2 | 83.0 | 0.323 | 65.7 | 86.2 | 38.4 |
| Mistral (mistral-large-2512) | 71.8 | 88.6 | 82.7 | 0.317 | 65.7 | 85.8 | 3.8 |
| Anthropic (claude-sonnet-4-5-20250929) | 70.5 | 98.1 | 82.6 | 0.279 | 71.8 | 89.4 | 39.4 |
| Anthropic (claude-haiku-4-5-20251001) | 70.5 | 98.1 | 82.6 | 0.279 | 71.8 | 89.4 | 38.3 |
| Qwen (qwen-2.5-72b-instruct) | 69.5 | 92.0 | 81.5 | 0.241 | 74.6 | 89.8 | 2.8 |

Table 2. Per-model derived metrics and operational profile for all 40 models on the full 492-scenario benchmark.
| Model | Precision (%) | NPV (%) | F1 (%) | Youden's J | FPR (%) | Alert Rate (%) | Median RT (s) |
| --- | --- | --- | --- | --- | --- | --- | --- |
| GLM (glm-5.1) | 68.7 | 100.0 | 81.4 | 0.215 | 78.5 | 92.1 | 42.1 |
| Google (gemini-2.5-pro) | 68.2 | 100.0 | 81.1 | 0.199 | 80.1 | 92.7 | 21.3 |
| OpenAI (gpt-4o-mini) | 69.0 | 74.2 | 79.8 | 0.216 | 72.9 | 86.6 | 1.8 |
| Mistral (mistral-small-2603) | 64.7 | 100.0 | 78.5 | 0.061 | 93.9 | 97.8 | 1.6 |

Second, per-model confusion matrices (Figure M3) reveal that the per-cell counts vary in ways the aggregate single-metric report cannot reflect: high-recall models with comparable F1 differ substantially in their false-positive counts, and high-specificity models cluster around very different sensitivity values. The information lost in collapsing four cells into one composite score is what the three-tier framework recovers; the operational evidence supporting that claim is reported in the clinical paper (Proulx et al., in preparation).

**Figure M3.**
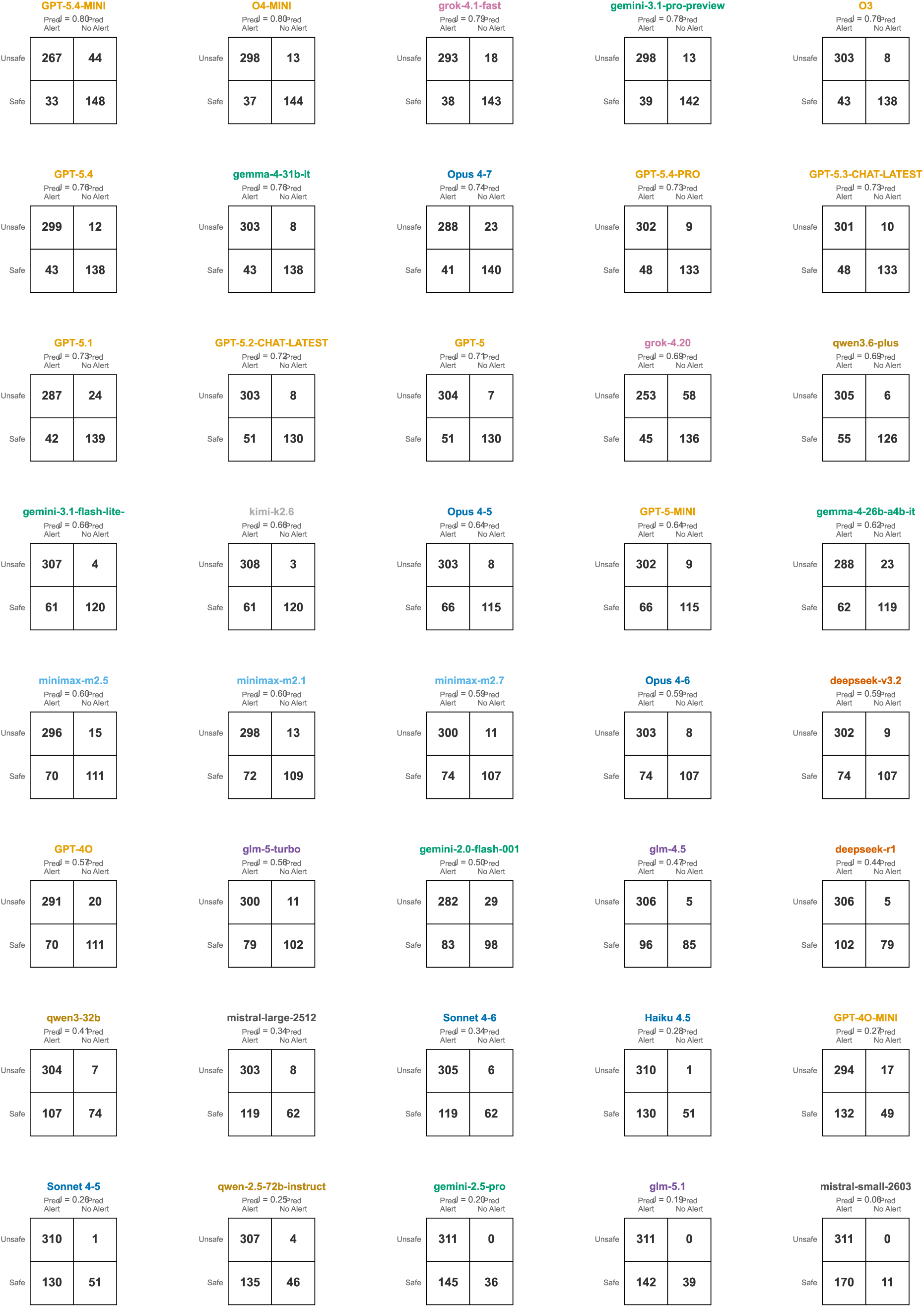
Confusion matrices for all 40 models on the full 492-scenario benchmark. Each 2×2 matrix shows true positive, false positive, true negative, and false negative counts from majority-vote classification. Rows are actual class (Unsafe / Safe); columns are predicted class (Pred Alert / Pred No Alert). Models are arranged in a 5-column grid sorted by Discrimination Youden’s J descending, with each model name shown above its matrix in the provider’s color and the Discrimination Youden’s J value below the model name. Cells are unfilled with a single thin black border and the count as the only in-cell signal, so the matrix reproduces cleanly in print and in grayscale.

## Discussion

We introduce PsiBench, a benchmark of 492 clinically validated medication-safety scenarios, and a non-overlapping three-tier evaluation framework that separates highest-stakes discrimination, the operational clinical decision support (CDS) regime, and category-correct alerting. The methodological contribution of this work is the framework and benchmark themselves; full empirical findings across 40 evaluated LLMs are reported in a separate clinical paper. The framework is general: any binary medication-safety classifier (rule-based, conventional ML, or LLM-driven) can be evaluated under these three tiers, and the scenario set has two decades of operational provenance in national hospital safety evaluation programs.

### Distinguishing operational benchmarks from academic benchmarks

A core methodological argument for PsiBench is the distinction between an academic benchmark and an operational benchmark. Existing LLM benchmarks in healthcare, including MedQA, Rx-LLM, and RxSafeBench, evaluate medical knowledge or pharmaceutical reasoning in academic contexts rather than operational medication safety.^30–34^ MedQA tests board-exam-style questions; Rx-LLM evaluates medication-related task performance across formulation matching, dose identification, and drug interactions; RxSafeBench assesses medication safety in simulated consultations. None reproduces the clinical workflow of a CPOE/EHR system evaluating a complete medication order against a patient’s clinical context, which is the task that hospital safety systems must perform thousands of times daily. A second methodological limitation compounds the first: most of these evaluations report a single aggregate metric (accuracy or F1) on an imbalanced scenario set, an evaluation design known to obscure systematic differences in sensitivity-specificity tradeoffs and to inflate apparent performance on the dominant class.^35–37^ Together, these two gaps mean that current LLM evaluations provide limited insight into whether these models could meaningfully improve medication safety in practice, for two reasons: they test the wrong task, and they report it in a way that hides the clinically decisive variation.

The PsiBench scenarios derive from annual, federally recognized audit criteria applied by more than 2,000 U.S. hospitals to certify medication-safety performance on their installed CPOE/EHR configurations.^15^ This is not a synthetic benchmark assembled for the LLM era; the scenarios encode the same clinical hazards that hospital safety systems are formally audited against every year, authored by the clinical pharmacology experts whose decades of experience underpin that annualized testing procedure. An LLM that performs well on PsiBench is, by construction, performing well on the exact task hospital CDS systems are certified to do.

### The three-tier framework

Standard LLM medication-safety evaluation uses a single binary metric (often F1 or accuracy) that collapses three fundamentally different clinical questions into one number. The three tiers each test a clinically distinct question on its own scenario subset, with a class mix chosen to make the corresponding metric informative.

The **Discrimination tier** isolates highest-stakes discrimination on a 98-scenario near-balanced subset of 50 fatal scenarios and 48 deception scenarios. Deception scenarios are safe medication orders intentionally constructed to resemble unsafe orders at first glance, with surface-level alarm features that resolve to a clinically appropriate order under closer reading. A naive always-alert classifier scores Youden’s J of 0 on this subset; the deception set provides a tight adversarial specificity test that sharpens the question to the discriminative reasoning the tier is meant to probe.

The **Operational tier** uses the 394-scenario class mix a deployed system must handle: 261 serious unsafe orders against 133 safe orders (92 regular safes plus 41 Excessive Alerts reclassified as operational negatives). The reclassification is the methodologically important move: Excessive Alerts scenarios are contexts where rule-based CDS systems are known to over-alert, and a capable safety model should refrain rather than fire. Scoring restraint on this subset approximates the alert-fatigue dynamics of production use.

The **Attribution tier** evaluates an orthogonal dimension: whether the model alerts for the correct clinical reason. For each correctly triggered alert on an alert-required scenario (311 in total), the predicted safety-category array is compared to ground truth by set-membership matching. Attribution is independent of detection quality; a model can have excellent discrimination and still attribute alerts to the wrong category, and reporting attribution alongside detection is the contribution of this tier.

The Discrimination and Operational tiers are non-overlapping, and together they cover all 492 benchmark scenarios. The Attribution tier evaluates an orthogonal property across the 311 alert-required scenarios. Each scenario is tested exactly once with the methodology most appropriate to its severity and class role.

### Operational deployment context for the framework

The clinical-deployment context for this framework, including the regulatory and clinical-operations arguments for why operational-grade evaluation matters at all, is developed in a separate clinical paper. From a methodology standpoint the requirement is straightforward: tier-specific metrics tied to tier-specific clinical questions, scenario provenance grounded in national hospital Health IT audit experience, and a non-overlapping scenario partition that prevents trivial scenarios from inflating headline metrics.

### Limitations of the framework

PsiBench is 492 scenarios drawn from 11 categories; it does not exhaust the long tail of medication errors observable in production CPOE/EHR systems. Three runs per model at temperature 0.2 capture central-tendency behavior but underestimate the tails of the response distribution. All prompts are in English; multilingual deployment may differ. The benchmark does not test adversarial prompts, retrieval-augmented context, or fine-tuning; reported performance reflects off-the-shelf capability via commercial APIs. Provenance limits also apply: the scenarios derive from an audit procedure focused on the highest-prevalence and highest-severity medication-safety failures, so the benchmark may be less informative for rare or emerging safety domains not represented in the audit set.

The deception subset is, by design, an adversarial specificity test rather than a sample of routine practice; specificity numbers on the Discrimination tier are not directly comparable to specificity numbers from observational CDS deployment data. The Attribution tier rests on the category taxonomy used by the audit procedure, and some scenarios admit more than one defensible categorization; the set-membership matching rule is the appropriate response to this taxonomy ambiguity, but it does not eliminate it.

### Forward-looking applications of the framework

PsiBench is necessary but not sufficient for clinical deployment. A high tier score demonstrates capability on a curated set of scenarios; it does not guarantee performance on the patient population or formulary of a specific hospital. Real deployment requires prospective validation against the local workflow and ongoing post-market surveillance. The three-tier framework provides an evaluation structure for that continuing process: the Discrimination tier monitors regression on the highest-stakes discrimination capability, the Operational tier tracks operational calibration, and the Attribution tier audits reasoning attribution. Periodic re-evaluation against the framework, with the scenario set updated annually in step with the national audit procedure, would surface model regression or drift that an installation-time evaluation alone could miss.

The framework is also extensible. Ensemble architectures that pair a Discrimination leader, an Operational leader, and an Attribution leader can be tested against any single model on the same three tiers, and the same rubric applies to non-LLM classifiers, enabling head-to-head comparison of LLM, rule-based, and hybrid CDS approaches.

## Methods

### Benchmark Design and Clinical Provenance

PsiBench is a medication ordering safety benchmark comprising clinically validated scenarios created by clinical pharmacology experts with decades of experience in hospital medication safety evaluation. This expertise underpins an annualized testing procedure used to evaluate CPOE/EHR systems in more than 2,000 U.S. hospitals.^40^ Each test case defines a clinical scenario containing a test patient with a specific medical context (diagnoses, current medications, laboratory values, allergies, demographic information) and a medication order to be evaluated. The task for each scenario is binary classification: determine whether the medication order requires a safety alert (unsafe) or may proceed without intervention (safe). When alerting, models are additionally asked to cite the applicable safety category or categories. This mirrors the core function of CPOE/EHR clinical decision support systems in production hospital environments.

The benchmark defines “unsafe orders” with a deliberately high bar: medication orders known to cause fatal or severe patient harm, not merely potentially unsafe or theoretically problematic prescriptions. The evaluation prompt frames the task as evaluating a hospital’s clinical decision support system, asking the model to review a medication order against the patient’s clinical information and determine what should happen when the order is placed. This framing grounds model responses in the same decision framework used by hospital systems rather than abstract safety judgments.

### Scenario Composition and Coding

The benchmark comprises 492 unique clinical scenarios distributed across 11 safety categories and 3 subcategories (Figure 1). The categories are: Drug Interaction (79 scenarios), Excessive Dosing (100), Drug Dose Daily (55), Drug Laboratory (51), Drug Diagnosis (42), Drug Dose Single (42), Excessive Alerts (41), Drug-Allergy (23), Drug Route (21), Drug Age (20), and Drug Monitoring (18). The three subcategories are Drug-Drug Interaction, Therapeutic Duplication, and Renal. Each scenario is coded with four analytically relevant fields:

- **Severity** (fatal, serious, none). 50 scenarios are fatal, 261 are serious, and 181 carry no expected harm.
- **Ground truth alert decision.** Each scenario is labeled with a binary expected outcome: an unsafe order requires an alert; a safe order does not.
- **Deception flag.** 48 safe scenarios are authored to look dangerous at first glance so that specificity can be tested against an adversarial negative class rather than trivially safe orders. The deception flag is test-design metadata; it is recorded on the scenario but is never exposed to the model.
- **Category and subcategory labels.** Each scenario is labeled with the safety category that makes the order unsafe (or, for safe orders, the category the order nominally touches).

The 41 Excessive Alerts scenarios are reclassified as operational negatives (an alert would constitute an over-alert; we therefore expect “no alert” from a capable model). Rationale: Excessive Alerts scenarios are cases where rule-based CDS systems are known to over-alert in real deployment, and a capable safety model should recognize them as contexts where alerting would contribute to alert fatigue rather than patient safety. Treating them as negatives in the operational tier tests exactly this capability.

### Non-Overlapping Three-Tier Evaluation Framework

Standard LLM medication safety evaluation uses a single binary metric that collapses three fundamentally different clinical questions into one number. We propose a non-overlapping three-tier framework that tests each question on its own scenario subset.

#### Discrimination tier (highest-stakes discrimination)

Discrimination comprises the 50 fatal-severity scenarios plus the 48 deception scenarios, yielding 98 scenarios total. The class mix is near-balanced at 51.0% fatal and 49.0% deception. The naive always-alert baseline scores 51.0% accuracy and Youden’s J of 0 on this subset, so the full binary confusion matrix (sensitivity, specificity, Youden’s J, F1) is genuinely informative. Discrimination tests whether a model can distinguish lethal orders from safe orders specifically designed to resemble them.

#### Operational tier (operational CDS regime)

Operational comprises the 261 serious-severity unsafe orders plus the 133 non-deception safe orders, yielding 394 scenarios total. The 133 negative scenarios comprise 92 regular safe orders plus 41 Excessive Alerts scenarios reclassified as negatives. The class mix (66.2% serious unsafe, 33.8% safe) approximates the operational CDS regime a deployed system must handle. Full binary confusion matrix is computed.

#### Attribution tier (category-correct alerting)

Attribution comprises the 311 alert-required scenarios (50 fatal + 261 serious). Excessive Alerts scenarios are excluded because they are negatives in the operational framework. For each correctly triggered alert on an Attribution-tier scenario, the model’s predicted safety categories (returned as an array of category codes in the structured JSON response) are compared against the ground truth category using set-membership matching: a category match is recorded if the ground truth category appears anywhere in the model’s predicted category array. Category labels are normalized (full names, code abbreviations, and appended descriptions are mapped to the canonical category name) before comparison.

Non-overlapping property: Discrimination (98 scenarios) and Operational (394 scenarios) together account for all 492 benchmark scenarios with zero overlap. Each scenario is tested exactly once with the methodology most appropriate to its severity and class role. Attribution evaluates an orthogonal dimension (category correctness) across the 311 alert-required scenarios (50 fatal from the Discrimination tier plus 261 serious from Operational).

### Clinical Scenario Presentation and Prompt Design

Each scenario is presented to the model as a structured clinical vignette containing patient demographics, active diagnoses, current medication list, relevant laboratory values, known allergies, and the medication order under evaluation. The evaluation prompt instructs the model to act as a hospital CDS system evaluator and to determine what should happen when the order is placed: the order proceeds without intervention, or the order does not proceed cleanly and a safety alert is warranted (including cases where the order cannot be entered as written). Any outcome other than the order being entered and signed with no advice or information returned is treated as an alert; only a cleanly entered order is treated as no-alert. When the model indicates an alert, it must identify all applicable safety categories using standardized codes and provide brief clinical reasoning for each. The response is returned as a structured object containing the alert decision, an array of category-reasoning pairs, and any applicable subcategory codes. All models receive identical scenario text and instructions; no few-shot examples, retrieval-augmented context, or external drug database access is provided.

### Model Selection

We evaluated 40 frontier LLMs representing the current state of the art across diverse architectures and providers (Table 1). Models span 10 providers across the United States, China, and France: OpenAI (12 models), Anthropic (6), Google (6), Alibaba (3), MiniMax (3), Zhipu (3), xAI (2), DeepSeek (2), Mistral (2), and Moonshot (1). Selection criteria included commercial API availability, diverse architectures and parameter scales, coverage of both conservative and aggressive alerting profiles, and inclusion of reasoning-tier and conventional models from each major provider where available. Models were accessed through their respective commercial APIs or, where direct access was unavailable, through the OpenRouter API gateway.

### Evaluation Protocol

Each of the 492 scenarios was evaluated 3 times per model at temperature 0.2 (or the provider default where temperature is not configurable), yielding 59,040 individual assessments (492 x 3 x 40). Evaluations were conducted between April 21 and 23, 2026. For each evaluation, the model received the complete clinical scenario and returned a structured response. Responses were parsed programmatically into the binary alert decision and the category predictions. Responses that could not be parsed, or that returned an error, were treated as a failure of the model to handle the scenario safely and were scored as incorrect, rather than defaulted to a no-alert (which could be spuriously credited as a true negative on a safe order) or to an alert.

### Performance Metrics

Scenario-level metrics use majority-vote aggregation across the 3 runs per scenario: the final classification is the majority prediction (at least 2 of 3 runs agreeing). This reduces the influence of stochastic variation while preserving information about consistency. We report accuracy, precision (PPV), recall (sensitivity), specificity, F1 score, false positive rate (FPR = 1 - specificity), alert rate, Youden’s J (sensitivity + specificity - 1), and negative predictive value (NPV). NPV is sensitive to the positive base rate of the evaluated subset: a naive always-safe classifier achieves NPV equal to the safe base rate of the subset, so NPV claims must always be interpreted against this baseline. Category match is computed, across each of the 40 models, as the proportion of correctly triggered alerts on the Attribution tier scenarios where the predicted category array contains the ground truth category under set-membership matching.

### Consistency Analysis

For each model and scenario, we classified the three-run outcome as always correct, always incorrect, or inconsistent (mixed results). These three classes are exhaustive and mutually exclusive. We report per-model rates across all 492 scenarios. High consistency does not imply correctness: always-incorrect scenarios reflect stable but systematically wrong predictions, which are clinically more dangerous than inconsistent ones because they do not surface in internal reliability checks.

### Response Time Measurement

Response time was measured as wall-clock duration from API request submission to complete response receipt, including network latency and provider-side queuing. We report median per-model response time in Table 2.

### Data Management and Reproducibility

All benchmark data is stored in a relational database managed by a purpose-built Python package. The schema records each model, each safety category and subcategory, each clinical scenario (with its severity classification, expected alert decision, deception flag, category and subcategory labels, and a masked publication identifier), each per-run evaluation result (with the model’s alert prediction, category-match flag, response time, and token usage), per-scenario consistency aggregates across the three runs, and per-model overall aggregates. Medication text and model reasoning text are held in private tables that are never included in published exports.

To prevent benchmark contamination, individual scenario content (patient vignettes, medication names, specific clinical parameters) is not published. Public data uses masked category-prefixed labels (for example DRUG_ALLERGY_003) rather than clinical identifiers. Only aggregate performance statistics, category-level metrics, and masked scenario identifiers are included in the publication exports. Ground-truth severity labels and the deception flag are design metadata and are not exposed to the model at evaluation time.

### Software

The evaluation pipeline, data management layer, scoring methodology, and figure generation code are publicly available at the repository referenced in the Code Availability Statement. All figures and tables are generated from Jupyter notebooks that query the database directly, ensuring reproducibility of every reported number from the provided aggregate data.

## Data Availability

The benchmark scenarios and the per-scenario, per-model raw evaluation results are not released, in order to preserve benchmark integrity and prevent exposure of scenario content to LLM training corpora. The aggregate per-model performance data underlying the figures and tables in this manuscript will be made available upon publication in a form that preserves the confidentiality of the benchmark scenarios. Requests for access for replication or methodological extension may be directed to the corresponding author.

## Code Availability

The code used to generate the figures and tables in this manuscript will be made available upon publication in a form that preserves the confidentiality of the benchmark scenarios. The evaluation harness and scoring code that would expose benchmark content remain held privately.

## Ethics

This study evaluates LLM performance on standardized medication safety test cases. The benchmark contains no protected health information, no real patient records, and no clinical encounter data. All clinical scenarios are synthetic test cases created by clinical pharmacology experts using synthetic test patients designed for system evaluation purposes. No institutional review board approval was required. The study did not involve human subjects.

## Author Contributions

CRediT roles below are populated from the author intake form.

**Joshua Proulx:** Conceptualization; Methodology; Software; Validation; Investigation; Resources; Writing – review & editing.
**Bryce Daines:** Conceptualization; Methodology; Software; Validation; Formal analysis; Investigation; Writing – original draft; Writing – review & editing; Visualization.
**Michael Barton:** Validation; Writing – review & editing.
**Molly E. Leonard:** Validation; Writing – review & editing.
**Joseph A. Garcia:** Writing – review & editing.
**Bronson Young:** Validation; Writing – review & editing.
**Quinn Snell:** Writing – review & editing.
**Timothy W. West:** Writing – review & editing.
**Sam R. Watson:** Writing – review & editing.
**Maryam AlQaseer:** Writing – review & editing.
**Mathieu Louiset:** Writing – review & editing.
**Muhammad Bilal Maqsood:** Writing – review & editing.
**Mary J. Voutt-Goos:** Writing – review & editing.
**Caryn Douma:** Writing – review & editing.
**Nishaminy Kasbekar:** Writing – review & editing.
**Jaclyn Jeffries:** Writing – review & editing.
**Wadie Abu-Rahmeh:** Writing – review & editing.
**Karen Frush:** Writing – review & editing.
**Darshan K. Grewal:** Writing – review & editing.
**Mouna Bahsoun:** Writing – review & editing.
**Michael Leonard:** Writing – review & editing.
**Allan Frankel:** Writing – review & editing.
**David C. Classen:** Conceptualization; Writing – review & editing; Supervision.
**Stanley L. Pestotnik:** Methodology; Validation; Data curation; Supervision.

All listed authors have confirmed ICMJE authorship consent for this manuscript.

